# Lattice Model of Mitigated Epidemic

**DOI:** 10.1101/2020.06.15.20132191

**Authors:** Dmitry A. Garanin, Eugene M. Chudnovsky

**Affiliations:** Physics Department, Herbert H. Lehman College and Graduate School, The City University of New York, 250 Bedford Park Boulevard West, Bronx, New York 10468-1589, USA

## Abstract

We study a statistical lattice model of a mitigated epidemic. The level of mitigation, defined by measures to slow down the spread of the infection, is characterized by the infection transmissivity. It is determined by people’s mobility, frequency of contacts, and probability to catch the virus from a contact. In the absence of testing the infected people are isolated for a finite period of time during which they are symptomatic. In the presence of testing, people become isolated a soon as they are tested positive. We compute time dependence of daily new infections as function of transmissivity, initial infection, and testing. The duration of the epidemic increases rapidly with the increased level of mitigation while the number of people falling sick daily decreases. Testing, regardless of the level, has little effect on the duration of the epidemic. The total number of people who contract the disease over the lifetime of the epidemic depends weakly on its duration. It does not change significantly for the homogeneous testing of the population at the level below 10% daily.

## I. INTRODUCTION

Mathematical modelling of epidemics started with a seminal work of Bernoulli [1]. Modern mathematical epidemiology grew out of three articles published by Kermack and McKendric [2]. The century that followed has produced countless articles and books on the subject, see, e.g., Ref. 3 for a recent review and a recent book, Ref. 4. The topic has grown into the field of epidemiology that combines practice of medicine with applied mathematics. In recent months it received an increased attention due to the global pandemic of COVID-19. Inspired by the urgent societal need and by the appeal [5] of the federal funding agencies to contribute ideas that may help address COVID-19, physicists [6] and engineers [7] began applying skills acquired from modelling of many-body systems and computer networks to the description of the pandemic.

In this paper we study a statistical lattice model of the dynamics of COVID-19 that incorporates features of its outbreak and mitigation. We do not pretend to compete with comprehensive computer models developed by epidemiologists. Our limited goal is to understand in simple terms how the decrease in the transmissivity of the virus (achieved through lockdown, social distancing, face masks, etc.) and testing affect the severity and the duration of the epidemic, as well as the prospect of its management by the health system.

The model is described in Section II. The results of the simulations are presented in Section III. Our conclusions are formulated in Section IV.

## II. THE MODEL

We are implementing a model of the epidemic on a square lattice of size *N*_*x*_ × *N*_*y*_ up to 500 × 500. The number of lattice sites coincides with the size of the population. All persons are initially allowed to make *N*_*v*_ random visits per day to neighboring sites in the range *d*_*v*_. Probability of the visit, *p*_*v*_, is controlled by measures designed to reduce the spread of the infection. Normal funcioning of the population corresponds to *p*_*v*_ = 1 while total lockdown corresponds to *p*_*v*_ = 0.

In the initial state, most of the persons do not carry the virus and are labeled by the Green color. Small fraction *c*_*i*_ of the persons are initially infected and labeled by the Red color. If a Red visits a site occupied by the Green, or a Green visits a site occupied by the Red, the Green becomes infected and converts to Red with a probability *p* that accounts for personal protection.

Infected Red are initially asymptomatic. They continue to spread infection during the incubation period of *N*_*a*_ days, after which they become symptomatic and convert to Brown. The Brown are self-quarantined or hospitalized. They do not spread the infection. After the period of *N*_*s*_ days the Brown recover, become immune, and convert to Blue.

To account for testing we introduce a number of tests, *c*_*t*_, per day per person. Testing level, e.g., *c*_*t*_ = 0.1 means that 10% of the population are tested every day. Only the Green and the Red are tested because conditions of the Brown and Blue are considered to be known. Once the Red is tested it converts to Brown. If the fraction of Green and Red falls below *c*_*t*_, all of them get tested in one day and quarantined if needed, which stops the spread of the infection.

The overall progression of the epidemic is characterized in our model by the parameter *pp*_*v*_*N*_*v*_ that we call transmissivity, and also by *d*_*v*_, *N*_*a*_, and *c*_*t*_. In simulations presented below we choose *N*_*a*_ = 5, *N*_*s*_ = 20, *N*_*v*_ = 10, *d*_*v*_ = 1, and vary other parameters.

Depending on these parameters, there are sub-viral and viral regimes. In the sub-viral regime the infection may flare out locally but becomes quickly suppressed, with the Green remaining in the majority at the end. In the developed viral regime the Green are in minority when the epidemic ends. We compute the boundary between the two regimes numerically and show that it agrees with the condition on the virus replication rate, *R*_0_ = 1, used by epidemiologists. The transition between the two regimes resembles a non-equilibrium phase transition.

The spread of the infection in our model is similar to the flame propagation. In the initial state, there are burning hotspots far away from each other. In the sub-viral regime, they become extinguished. The final number of sick is very small and depends on the initial infection. In the viral regime, fluctuating burning fronts grow from each hotspot independently. The speed of the propagation of the front of the infection increases with transmissivity. The fronts propagating from individual hotspots eventually meet. The process continues until almost all “flammable” material (Green) gets burned (converts to Brown) and all Brown converts to Blue, or when the “flame” becomes extinguished by low transmissivity.

In fighting the infection the goal must be to achieve a sub-viral regime. However, for highly contagious diseases, such as seasonal flu and COVID-19, this goal is difficult to achieve. For that reason, while providing the conditions needed to achieve the sub-viral regime, our focus is on the viral regime that allows epidemic to take its course. Factors such as the weakening of the virus and/or vaccination, that can stop the spread of the infection, can be taken into account by simply making everyone Blue when it happens. At that point in time all the computed curves describing daily infections would drop down to zero. We are not considering such a scenario here.

## III. RESULTS

The spread of the infection from one infected person, illustrating the flame analogy, is shown in Fig. 1. It starts as a growing area of Red (infected, asymptomatic). After the incubation period of *N*_*a*_ = 5 days, the Brown area of quarantined people develops inside the Red area. Red and Brown fronts continue to expand until the Brown who fell ill first begin to recover and turn Blue. At that time, determined by the recovery period of *N*_*a*_ = 20 days, the Blue area develops inside the Brown area. Areas of all colors continue to expand, with Blue inside Brown inside Red, until the Blue occupies almost the entire region. The spread of the infection that begins from multiple sources is shown in Fig. 2.

**Figure 1:**
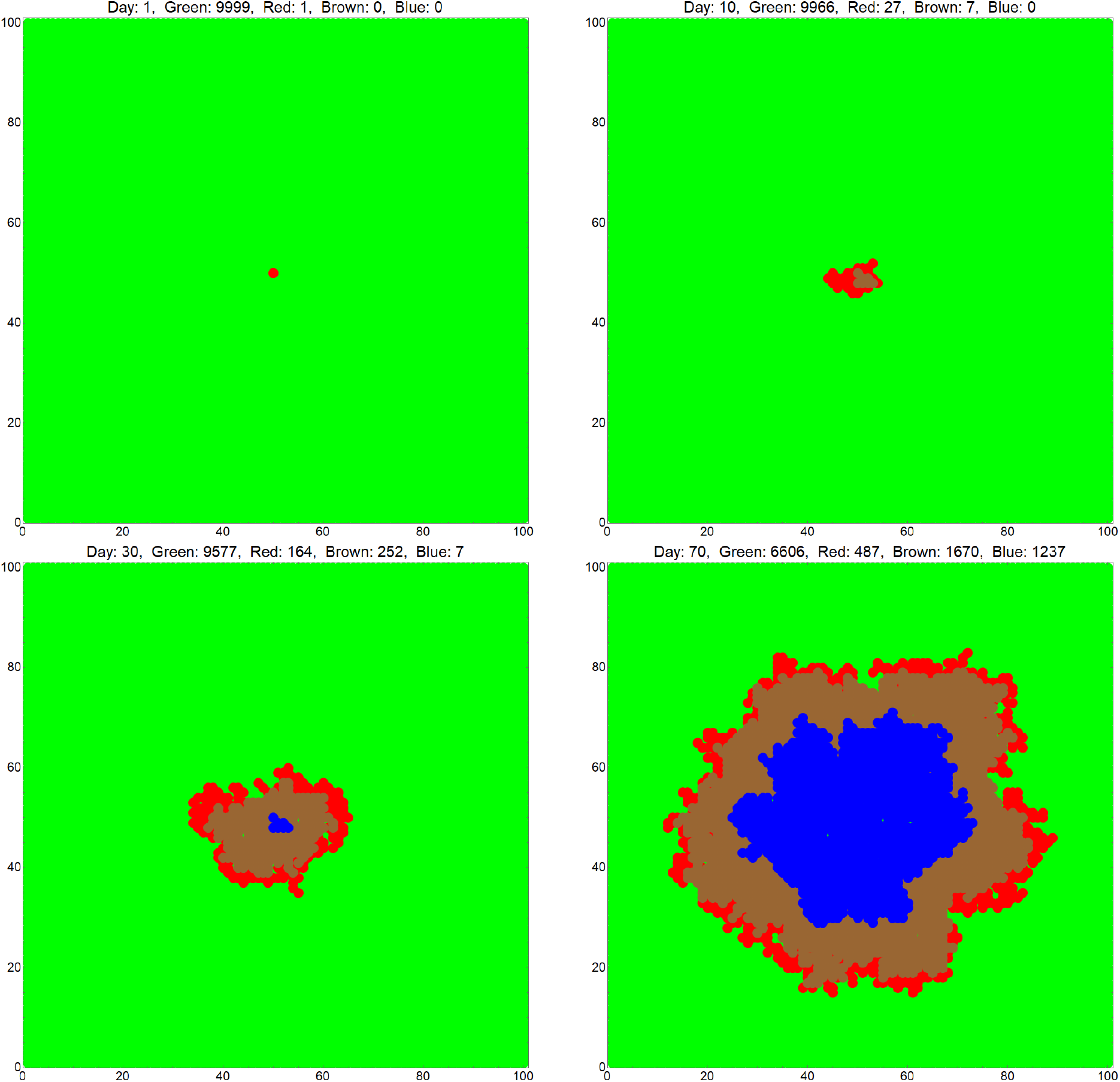
Progression of the epidemic started by one spreader. Green: Never Infected, Red: Infected-Asymptomatic, Brown: Symptomatic-Isolated, Blue: Recovered.

**Figure 2:**
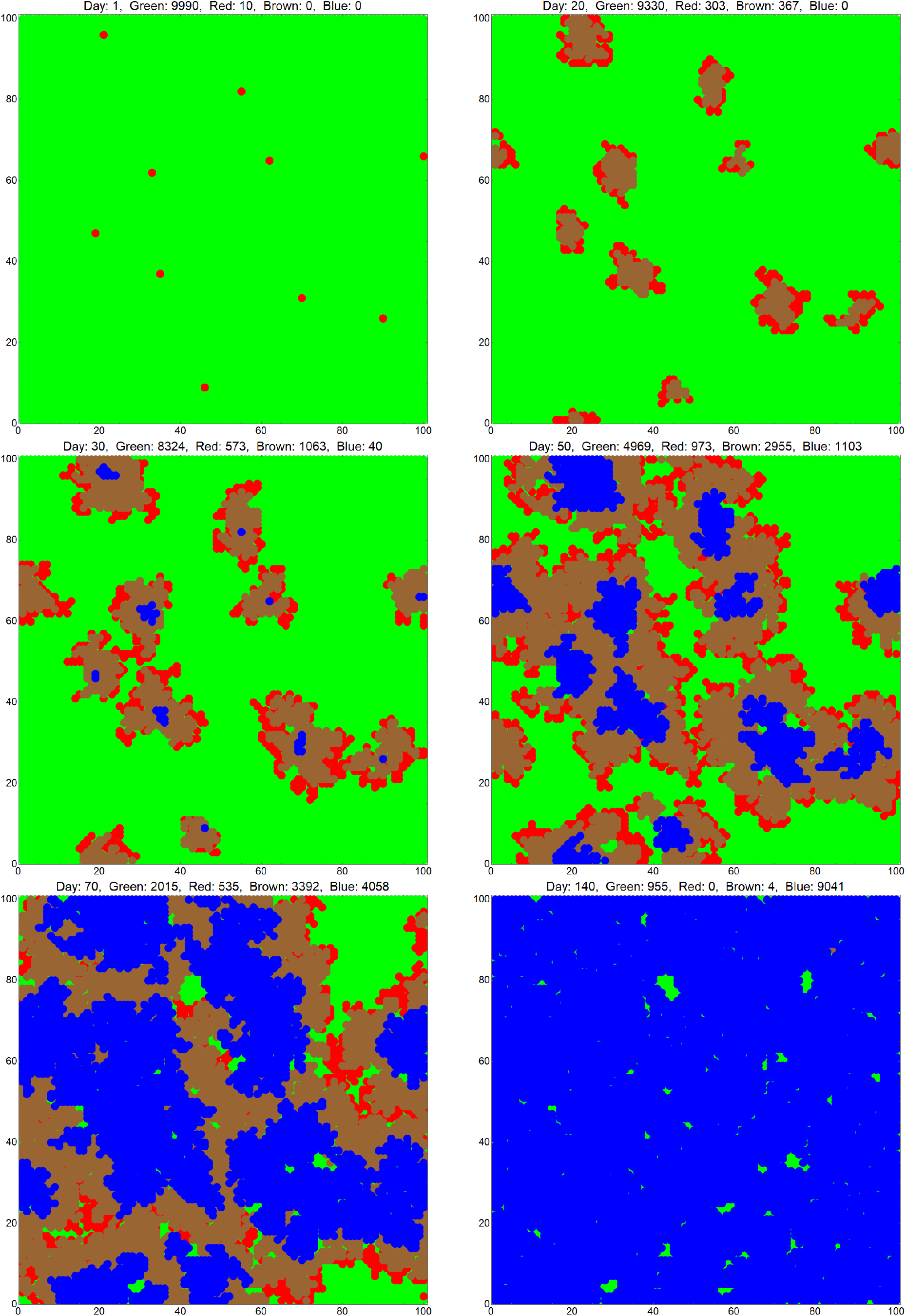
Progression of the epidemic started by multiple spreaders. Green: Never Infected, Red: Infected-Asymptomatic, Brown: Symptomatic-Isolated, Blue: Recovered.

As the epidemic progresses the parameter that concerns the health officials is the number of people who fall ill daily because 1) Most of them will try to contact their doctors and this may overwhelm capabilities of individual doctors to respond, and) A fraction of them will need hospitalization and the hospitals may run out of beds. Knowing that fraction, recovery and mortality rates, would allow one to easily calculate the total number of hospital beds required at any moment of time. Time dependence of the newly infected fraction of the population per day vs time is shown in Fig. 3 for the initial infection of 0.1% of the population and 0.01% of the population, for different levels of mitigation described by the transmissivity *pp*_*v*_*N*_*v*_. This computation does not include testing (*c*_*t*_ = 0). Few observations are in order. The duration of the epidemic increases rapidly with the increased level of mitigation. The number of daily new sick goes down accordingly. However, the total number of people who fall sick during the lifetime of the epidemic uninterrupted by weakening of the virus and/or immunization shows weak dependence on its duration.

**Figure 3:**
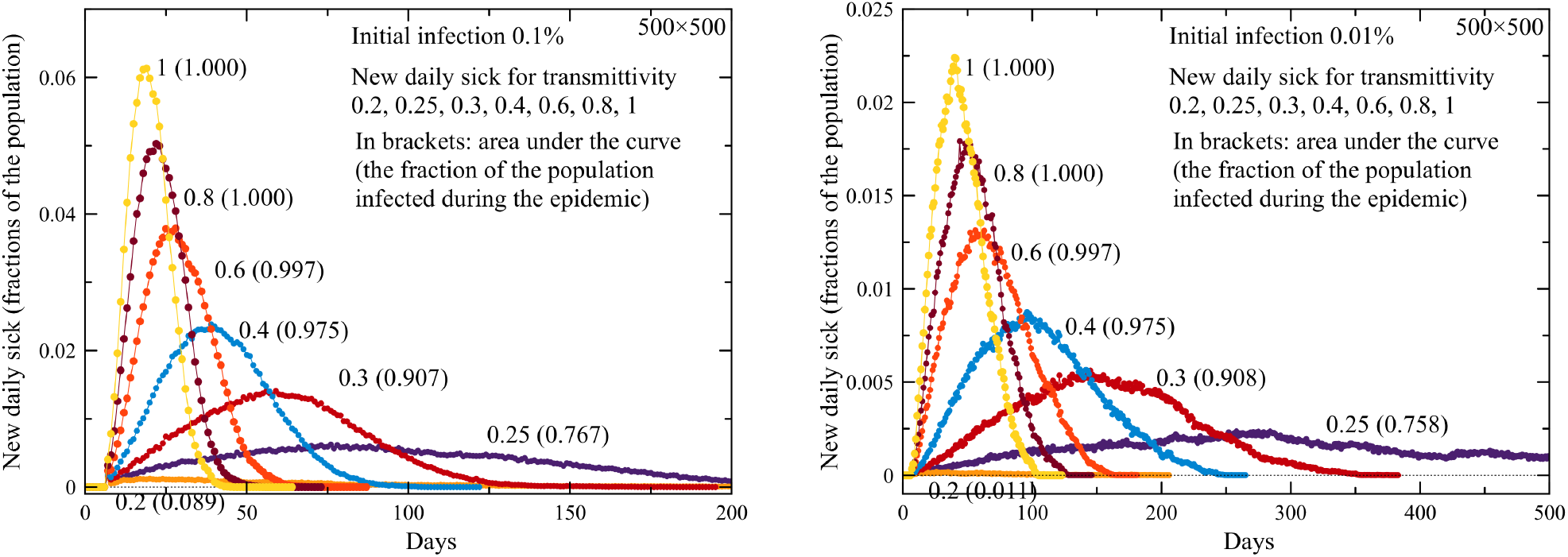
Daily new sick vs time for different transmissivities. Left: 0.1% of the population infected initially. Right: 0.01% of the population infected initially. The number in brackets shows the fraction of the population infected for the duration of the epidemic.

As Fig. 3 shows, the initial infection before the mitigation begins is an important factor for the duration of the epidemic. It is roughly triples when the initial infection changes from 0.1% (Fig. 3-left) to 0.01% of the population (Fig. 3-right). The flame analogy explains why the duration of the epidemic becomes longer with the decreased level of the initial infection: It takes longer time for the independent “burning” regions to overlap. For the spread on a two-dimensional map, the duration of the epidemic increases roughly as 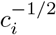, while the number of daily new sick decreases accordingly. Here we used *d*_*v*_ = 1. Allowing visits of distant sites during the epidemic, with *d*_*v*_ comparable to the size of the system, would make the epidemic faster and would result in the exponential growth of the number of newly infected people.

The next question we addressed was how the testing that isolates asymptomatic people, preventing them from spreading the infection, would change the dynamics of the epidemic. Here we consider homogeneous testing with no feedback on the mitigation by adjusting local transmissivity to the results of the testing. The latter would be a kind of an intelligent testing that can stop the epidemic in it tracks without allowing it to cross from sub-viral into the viral regime.

The dependence of the duration of the epidemic on transmissivity with and without testing is shown in Fig. 4-left. Testing moves the boundary between sub-viral and viral regimes towards higher transmissivity. However, the effect is significant only for a very large level of testing, like 10% of the population daily shown in the figure. It does not change the duration of the epidemic in the viral regime when the transmissivity is above 0.3. Such weak dependence of the duration on testing in the developed viral regime is confirmed by other choice of parameters.

**Figure 4:**
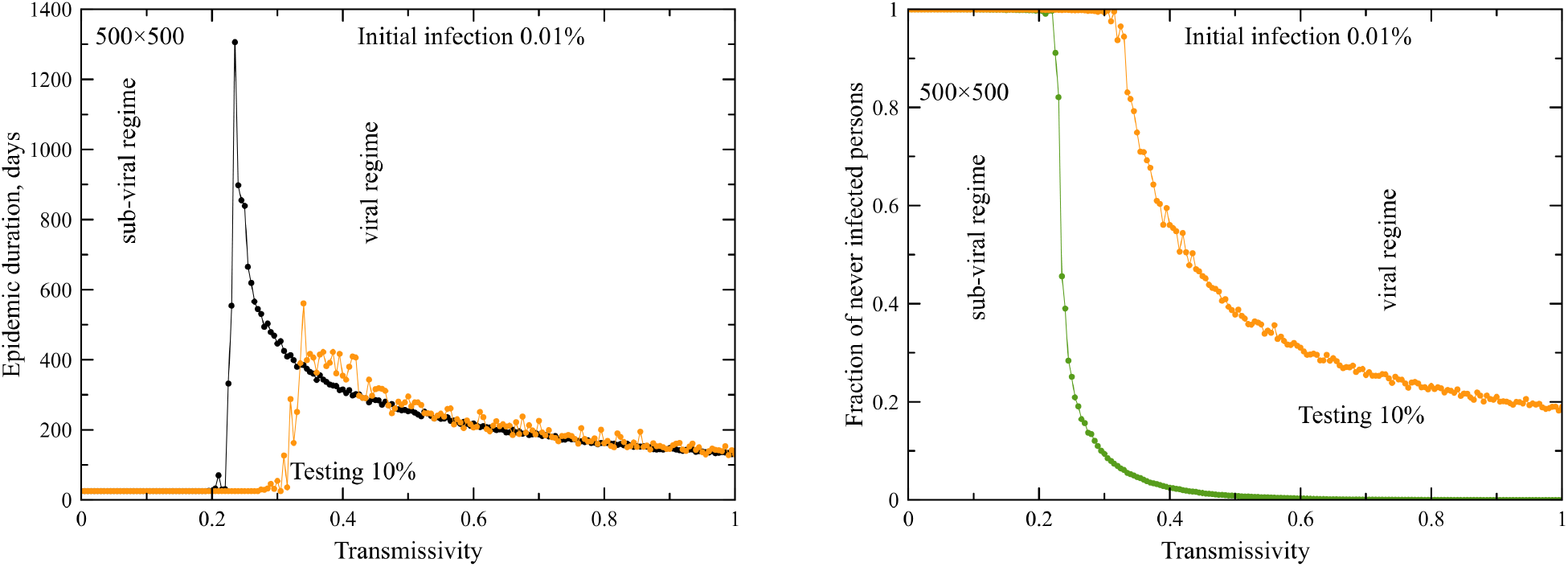
Effect of testing. Left: Duration of the epidemic vs transmissivity without testing (black) and with testing of 10% of the population daily (yellow). Right: Fraction of people that have never been infected over the course of the epidemic vs transmissivity without testing (green) and with 10% of the population tested daily (yellow).

On the contrary, the number of people that have never been infected during the course of the epidemic depends strongly on transmissivity, see Fig. 4-right. Without testing, it drops rapidly as one moves away from the boundary between sub-viral and viral regimes, resulting in the contraction of the disease by 100% of the population over the course of the epidemic. Homogeneous testing reduces that number but the effect, again, is significant only for a very large level of testing, like 10% of the population daily, which may be impractical.

In the absence of testing, the boundary between sub-viral and viral regimes can be estimated from the following argument. The Red making *p*_*v*_*N*_*v*_ visitations per day meet *N*_*a*_*p*_*v*_*N*_*v*_ Green in *N*_*a*_ days. With the probability to infect a Green being *p*, a Red infects on average *R*_0_ = *N*_*a*_*pp*_*v*_*N*_*v*_ Green before it turns Brown and gets isolated. Here *R*_0_ is the virus multiplication factor used by epidemiologists. The condition *R*_0_ = 1 provides the estimate for the critical transmissivity,

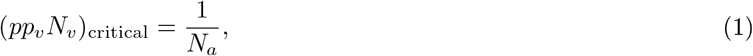

that separates sub-viral and viral regimes. For the incubation period *N*_*a*_ = 5 days used in our simulations it gives the critical transmissivity of about 0.2 which agrees with the simulations, see Fig. 4-left. In computer work we used for the visit distance *d*_*v*_ = 1. The dependence on *d*_*v*_ is weak. Increasing the visit distance from *d*_*v*_ = 1 to *d*_*v*_ = 10 results in a 30% reduction of the critical transmissivity.

The dependence of the number of people infected daily with and without testing is shown in Fig. 5 for transmissivity It confirms the findings represented in Fig. 4. The duration of the epidemic in the developed viral regime is not affected by testing. When the fraction of people tested daily is small, the effect of the testing on the number of people who contract the virus daily, is small too. To reduce that number by 50%, as much as 10% of the working population (∼ 15 million in the U.S.) would have to be tested daily.

**Figure 5:**
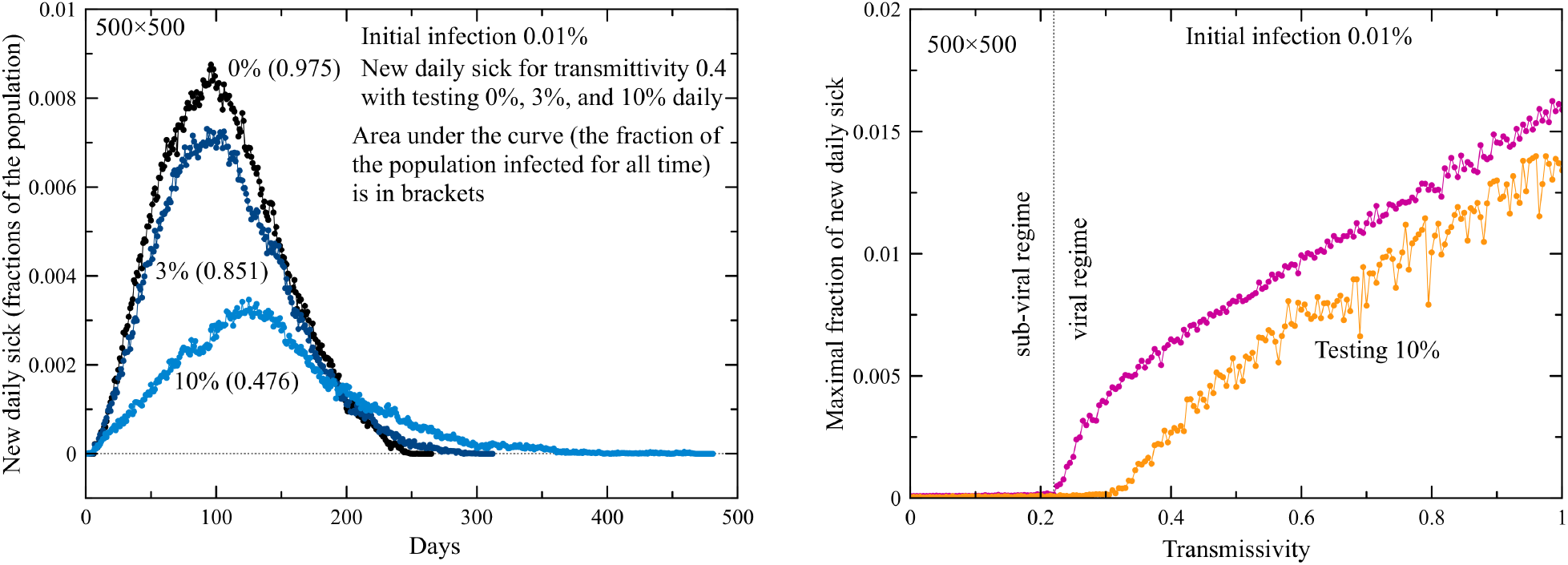
Effect of testing. Left: Daily new sick for transmissivity 0.4 and three different levels of testing: 0% of population tested daily, 3% and 10%. Right: Maximal fraction of daily new sick during the epidemic without testing (pink) and with 10% of the population tested daily (yellow).

## IV. CONCLUSIONS

- Early detection of the virus followed by a high level of mitigation through lockdown, social distancing, mandatory face masks, etc. is needed to keep the spread of the infection in a sub-viral regime that extinguishes quickly.
- Duration of the epidemic in the viral regime increases rapidly with the increased level of mitigation.
- Duration of the epidemic decreases rapidly with the number of infected people before mitigation begins.
- While the number of people infected daily goes down with the increased duration of the epidemic, the total number of people that contract the disease over the lifetime of the epidemic depends weakly on its duration.
- Testing, regardless of its level, has little effect on the duration of the epidemic.
- Homogeneous testing at the level below 10% of the population daily does not significantly affect the number of new daily infections or the total number of people that contract the disease during the lifetime of the epidemic. Intelligent testing of local population at a much smaller level, that results in the continuous adjustment of the mitigation in space and time, must be more effective.

In the light of the above findings and with account of the negative effect that the reduction in mobility has on the economic activity, education, health, etc., we conclude that, unless mitigation is aimed at stopping the infection from crossing to the viral regime (epidemic), or the intelligent testing is implemented in the viral regime, mitigation (such as lockdown, social distancing, wearing face masks, etc.) can only be justified by factors outside the parameter space that determines dynamics of the epidemic, such as

- Protecting the vulnerable (elderly and immune-compromised).
- Reducing the number of daily new infections to the level determined by the capabilities of the health system. Early detection of the epidemic is crucial in that respect.
- Stretching the lifetime of the epidemic in anticipation of the vaccine or seasonal changes that reduce the effectiveness of the virus.

## Data Availability

All the data related to this article are available from the authors.

## V. ACKNOWLEDGEMENTS

Authors’ research is supported by the U.S. Department of Energy, Office of Science, DOE Grant DE-FG0 - 93ER45487. Any opinions, findings, and conclusions or recommendations expressed in this material are those of the authors and do not necessarily reflect the views of the DOE.

